# Demonstrating reduced school absenteeism and disciplinary actions through a school-based teletherapy intervention among socioeconomically vulnerable youth: a retrospective observational study

**DOI:** 10.1101/2025.03.11.25323793

**Authors:** Akshay Swaminathan, Gloria Ye, Connor Weber, Gaurai Uddanwadiker, Alex Alvarado, Devika Bhushan

## Abstract

**Background:** Schools play a central role in addressing the mental health needs of young people. Here, we analyze data from a school-based teletherapy intervention delivered in two socioeconomically vulnerable school districts to characterize sociodemographics of referred students, identify variables associated with session completion, and assess associations with attendance, grade point average (GPA), and disciplinary actions.

**Methods:** Guidance counselors and mental healthcare professionals referred eligible students for participation in a 12-week teletherapy intervention in two socioeconomically vulnerable K-12 school districts (A and B) in California between 2021-2023. Using multivariable linear regressions, we assessed associations between session completion and attendance, GPA, and disciplinary actions during the school year following the intervention, as compared to the year prior.

**Results:** A total of 533 students were referred for the intervention in District A and 385 in District B. Of these, 389 (73%) and 258 (67%) attended an intake appointment; and 165 (31%) and 96 (25%) completed at least one session. In District B, completing 12 sessions was linked to an estimated increase of 12.31 school days attended (a 6.84% rise in attendance; 95% CI: 2.04% to 11.5%, p = 0.0051) and a reduction of 2.76 disciplinary actions, such as suspensions for behavioral incidents (95% CI: −0.48 to 5.88 fewer incidents, p = 0.091). In District A, students with a higher baseline GPA completed more sessions (1.3 additional sessions per 1-point higher GPA, 95% CI: 0.4 to 2.19, p<0.001). In both districts, students in foster care or experiencing homelessness completed fewer sessions.

**Conclusions:** This study demonstrates the potential of a school-based teletherapy intervention to boost attendance and reduce the need for disciplinary actions, particularly in socioeconomically vulnerable districts. Barriers to equitable participation, especially for students in foster care and experiencing housing instability, highlight the need for targeted support.

## Introduction

Youth mental health has been declared a national emergency (AAP, AACAP, & CHA, 2021) and a critical public health concern (U.S. Department of Health & Human Services, 2021), with rising rates of anxiety,depression, and isolation profoundly affecting students’ emotional well-being, connectedness, academic performance, and potential. Given that youth spend a significant amount of their day at school, schools play a critical role in addressing these challenges, often serving as the primary setting for identifying the need for and referring students to mental health services (Grande et al, 2023; Sekhar et al. 2021; Sowa et al, 2024). Despite this pivotal role, the evidence on the effectiveness of school-based teletherapy interventions in improving academic outcomes, including grade point average (GPA) and attendance, or for reducing disciplinary actions like suspensions and expulsions, remains limited. To our knowledge, quantitative data linking school-based teletherapy interventions to academic outcomes, attendance, and behavioral incidents for low socioeconomic status, recent immigrants, English language learners, and students with housing instability is limited and more targeted research is needed to establish definitive associations or causal relationships.

Untreated mental health conditions in school-age youth can lead to a cascade of negative outcomes for years to come, including diminished academic achievement, higher rates of absenteeism, and increased disciplinary actions like suspensions and expulsions (Finning et al., 2019; Kaushik, Kostaki, & Kyriakopoulos, 2016). Beyond the immediate school environment, these challenges can have long-term implications, such as reduced quality of life and limited future opportunities, such as diminished employment opportunities and lower incomes, as well as greater involvement with the justice system (Counts et al., 2025; Currie, 2024; Prins et al., 2022). In vulnerable populations—including students with low socioeconomic status, recent immigrants, English language learners, and students with housing instability—these consequences of untreated mental health conditions are amplified (Currie, 2024). Effective teletherapy interventions are essential to prevent these consequences and support students in reaching their full potential. Teletherapy interventions have great potential in increasing access to mental health care for youth, especially in underserved areas and for vulnerable populations (Chen et al., 2024; Grady et al., 2011; Spiegel et al., 2023). Between 2019 to 2002, there was an 830% increase in the number of Medicaid-enrolled youth aged 3 - 17 (Ali et al., 2023; Cummings et al., 2023), highlighting the demand for accessible healthcare among vulnerable youth. Notably, teletherapy interventions allow for more flexibility in access to care beyond the school day, offering students greater privacy and carrying less stigma, and without needing transportation or time off from work for an accompanying parent or caregiver (Cummings et al., 2023; Orsolini et al., 2021; Stephan et al., 2016).

School-based mental health interventions are typically categorized as either universal or targeted. Universal interventions are delivered to all students in a population, regardless of their mental health status, and are generally preventative in nature (Sekhar et al., 2021; Zhang et al., 2023). In contrast, targeted interventions focus on individuals identified as having elevated symptoms and are therapeutic, seeking to reduce symptoms and improve associated school outcomes (Zhang et al., 2023). Universal programs can also avoid stigmatization and reach broader populations, but are typically less effective at addressing the specific needs of high-risk individuals (Andrews and Foulkes 2025; Le et al., 2021; Lee et al., 2017; Zhang et al., 2023). While targeted approaches may be more effective than universal interventions in addressing specific mental health symptoms (Werner-Seidler et al., 2021), they also present unique challenges. Screening efforts required for targeted interventions can strain already tight resources in school settings, and the individualized nature of these interventions and their follow-up can lead to implementation challenges (Panchal, Cox, & Rudowitz, 2022; Zhang et al., 2023). Additionally, concerns about labeling and stigma associated with targeted programs may deter participation, particularly among vulnerable populations (Zhang et al., 2023). Given these trade-offs, there is a critical need to explore the implementation feasibility and effectiveness of targeted interventions in resource-constrained settings.

Here, we present a retrospective analysis of a school-based, targeted intervention that provided individualized teletherapy over 12 weeks, delivered by licensed therapists with pediatric behavioral health expertise. The intervention was implemented between 2021 and 2023 across two public school districts in California—both characterized by high levels of socioeconomic vulnerability and race/ethnicity diversity, including a high proportion of students from immigrant families. We analyze key process measures, including referral rates and session completion rates, and investigate the relationships between session completion and academic outcomes (GPA, disciplinary actions for behavioral incidents, and attendance). This study aims to provide actionable insights into the effectiveness of targeted school-based mental health interventions delivered by telehealth for academic success, and their potential to support vulnerable student populations.

## Methods

### Study design

This study utilized a retrospective observational design to analyze routinely collected data on a school-based, targeted teletherapy intervention (described below). We follow the STROBE checklist for reporting (STROBE Initiative, 2025). All students provided consent - a requirement for accessing care - to receive teletherapy from Daybreak Health. Since this study analyzed pre-existing data and introduced no additional risk to patients, the WCG IRB determined it does not qualify as human subjects research and did not require further review.

### Data sources

Data for this study were drawn from two primary sources: Daybreak Health Inc. and participating school districts. Daybreak Health provided data on student referrals, and session completion. Academic and sociodemographic data were obtained from the two participating school districts in California, which routinely collect information on student age, gender, race/ethnicity, school attendance, GPA, behavioral incidents, English language learner (ELL) status, Individualized Education Plan (IEP)/504 status, foster care, and homelessness status. All data were collected for the 2021-2022 and 2022-2023 academic years.

In addition to student-level data, district-level population indicators were obtained from the California School Dashboard. We round all district-level statistics to maintain anonymity. School District A has a total enrollment of >15,000 students, with more than 40% classified as socioeconomically disadvantaged (defined as being eligible for free or reduced lunch, or having parents/guardians who did not obtain a high school diploma), 40% identifying as Hispanic, and 10% identified as ELLs. School District B is a larger district with approximately 20,000 students, where over 90% of students are classified as socioeconomically disadvantaged, 80% identify as Hispanic, and over 20% identified as ELLs.

### Outcomes

The primary outcomes of interest included academic measures to assess the impact of the intervention. Academic outcomes included GPA recorded on a standard 4.0 scale (available for District A only); school attendance, defined as the proportion of instructional days attended during the academic year (available for both districts); and disciplinary actions for behavioral incidents, including disciplinary referrals, in-school suspensions, and expulsions (available for District B only). All outcomes were evaluated by comparing baseline data in the academic year before the intervention to follow-up data in the academic year after the intervention. Additionally, process measures were analyzed to evaluate student engagement with the intervention. These included referral rates, intake completion, and session completion.

### Intervention

The intervention consisted of a structured, individualized, evidence-based teletherapy program delivered by therapists employed by Daybreak Health. The program primarily employed cognitive behavioral therapy (CBT) techniques tailored for children and adolescents and was delivered in a teletherapy format. Students were referred to the intervention by school staff, typically in response to observed mental health or behavioral concerns. In District A, school counselors referred students directly to Daybreak Health, following standard school-based mental health referral procedures and Daybreak’s inclusion and exclusion criteria (below). In District B, however, referrals followed a modified process that incorporated an intermediary mental health team consisting of on-site therapists employed by the district. This team conducted an initial screening to assess student needs and determine whether the teletherapy intervention was appropriate or if in-house treatment was more suitable. Additionally, District B implemented a universal mental health assessment using a 65-question screener that evaluated students across multiple domains, including anxiety, depression, and trauma. This proactive screening approach allowed counselors to systematically identify students in need of mental health support, potentially influencing both the composition and level of need of the students ultimately referred to Daybreak.

### Inclusion and exclusion criteria

Referred students met the criteria for the Daybreak intervention if they were between the ages of 10 to 19 years in district A, and 5 to 19 years in district B, and presented with mild to moderate symptomology for disorders of mood, anxiety, executive functioning, identity development, adjustment to life events, emotional dysregulation, behavioral concerns and co-occurring substance use, learning, and/or pre-clinical disordered eating challenges. Students were excluded if they exhibited severe symptoms of the above, involving safety risks, eating disorders, substance use disorders, recent severe trauma, or Child Protective Services involvement. Students who met criteria were included in the analysis if they had been referred to the Daybreak Health program during the study period and had both baseline and follow-up data available for at least one academic outcome. Students were excluded if they had incomplete baseline data or were no longer enrolled in school during the follow-up period (SI Table 1).

**Table 1:**
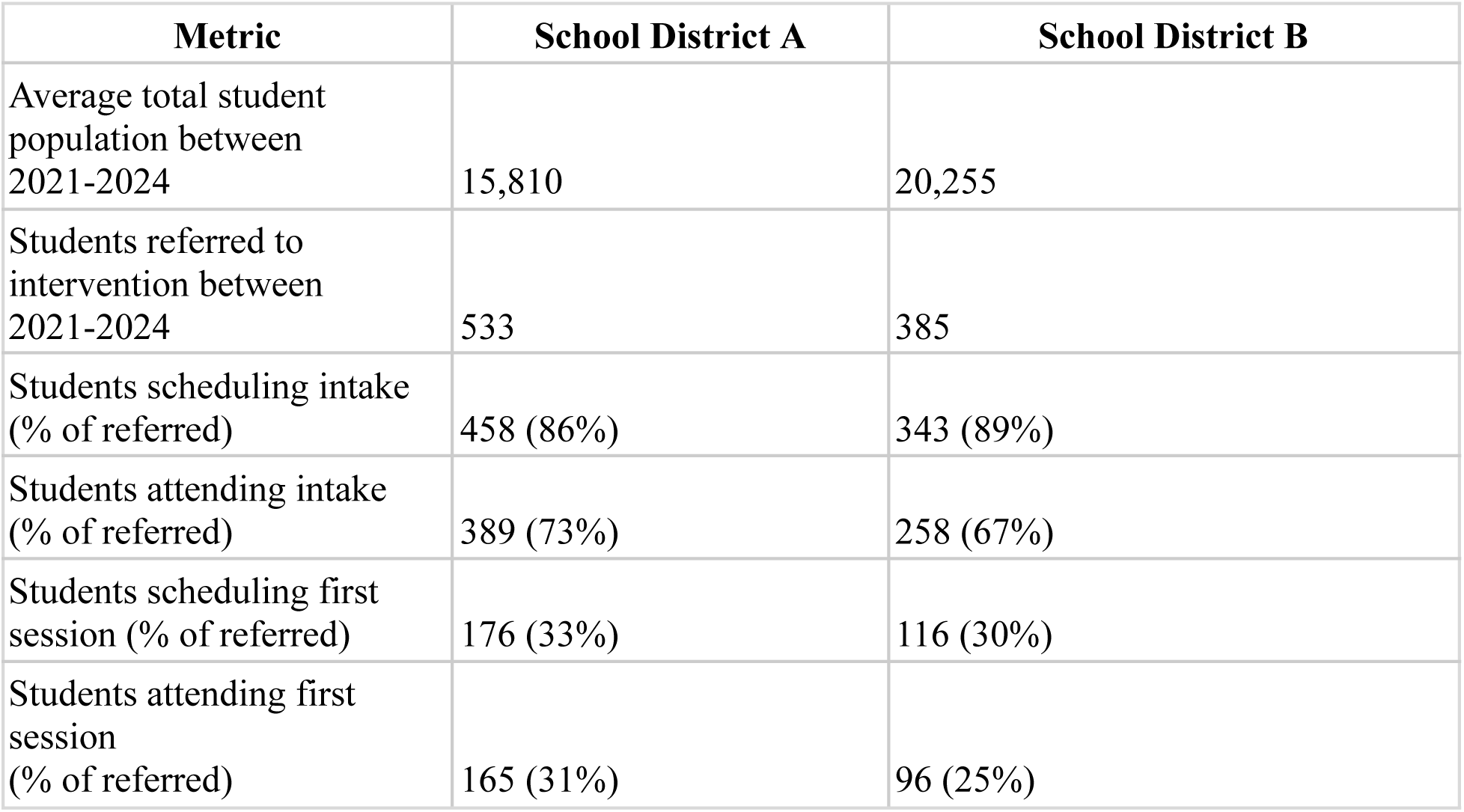
Overview of the student population and intervention referral funnel metrics for school districts A and B. Metrics represent the proportion of students advancing through each stage of the referral process: referred, scheduling an intake, attending the intake, scheduling a first session, and attending a first session. Percentages are calculated relative to the total number of students referred.

### Statistical analyses

We first summarized student demographic characteristics, referral metrics, and baseline academic measures using medians and interquartile ranges (IQR) for continuous variables, and counts and percentages for categorical variables. Differences between referred students and the overall student population were assessed to understand the characteristics of students who accessed the intervention. To assess student engagement with the intervention, we calculated the proportion of students progressing through each stage of the referral funnel, from referral to session attendance. This allowed us to quantify dropout rates at each stage and identify points of highest attrition, as well as demographic characteristics associated.

We used multivariable linear regression to examine the association between the number of sessions completed and changes in academic outcomes and disciplinary actions. Separate models were estimated for each outcome: yearly GPA, school attendance proportion, and disciplinary actions for behavioral incidents. To adjust for potential confounding, all models controlled for race/ethnicity, gender, English language learner (ELL) status, IEP/504 Plan status, foster care status, homelessness status, as well as the baseline values of the outcome variables.

To identify variables associated with sessions completed, we conducted additional multivariable regression models where the number of sessions attended served as the dependent variable. Independent variables included baseline attendance, GPA, ELL status, IEP/504 Plan status, foster care status, homelessness status, race/ethnicity, and gender. These analyses aimed to identify whether certain student characteristics were associated with greater or lesser engagement with the intervention. Missing data were addressed using median imputation for continuous variables, which was deemed appropriate given the relatively low proportion of missing values. Sensitivity analyses were performed to assess the robustness of findings to different imputation strategies, including mean imputation and multiple imputation methods. All analyses were performed using R (version 4.4.0).

## Results

### Student referral statistics

Between 2021 and 2024, 533 students were referred for care in District A, and 385 were referred in District B (Table 1). Among referred students, 86% in District A and 89% in District B scheduled an intake, with 73% and 67% attending the intake session, respectively. The stage with the highest attrition occurred between attending the intake and scheduling a first session, with 33% of referred students in District A and 30% in District B scheduling a first session. Ultimately, 31% of referred students in District A and 25% in District B attended a first session. These metrics provide insight into the progression of students through the referral process.

### Baseline characteristics

In School District A, 153 referred students who had complete GPA and attendance data were included in the final analysis. Approximately half of students were in grades 6–8 (46%) and the other half in grades 9–12 (54%). There were slightly more girls (66%) than boys (34%) among referred students. Compared to the overall district population, the referred group had a higher proportion of socioeconomically disadvantaged students (46% vs. 31% overall). Additionally, there was a notable representation of students with an Individualized Education Plan (IEP) or 504 Plan (18%). Baseline attendance among referred students was high, with a median attendance proportion of 95%. Of all students referred, 92 (60%) completed at least one session and 30 (20%) completed all 12. Compared to the overall School District population, referred students in District A were more likely to be socioeconomically disadvantaged (46%).

In School District B, 236 referred students that had complete behavioral incident data and attendance data were included in the final analysis. The referred population was predominantly younger, with most students in grades K–5 (39%) and 6–8 (51%), while only 9.7% were in grades 9–12. Similar to District A, referred students were slightly more likely to be female (69%). The referred group was overwhelmingly Hispanic (86%), reflecting the district’s overall demographic composition (80% Hispanic). The population of referred students also had a high prevalence of foster or homeless students (14%) and English language learners (ELL, 51%). Median attendance among referred students was 92%. These characteristics highlight the significant vulnerabilities and diverse needs of the referred population in District B. The district’s referred population was highly socioeconomically disadvantaged, with 98% of all students classified as such. Of all students referred, 129 (55%) completed at least one session and 28 (12%) completed all 12. Compared to the overall school district population, referred students in District B were more likely to be homeless or in foster care (4%) and be an English language learner (27%).

### Associations between session completion and academic outcomes

While some students in both districts completed multiple sessions, others participated minimally or did not attend any sessions after referral, highlighting variability in engagement with the intervention. In District A, session completion did not show notable associations with changes in academic outcomes. There was no association between sessions completed and post-intervention GPA (coefficient = 0.0024, 95% CI: −0.013–0.018, *p* = 0.76) or proportion of school days attended after the intervention relative to before (coefficient = −0.00047, 95% CI: −0.0021–0.0012, *p* = 0.58) (Table 3).

In District B, where there was a tiered approach to assigning students to Daybreak’s teletherapy intervention versus in-person care, session completion was significantly associated with improved school attendance rates. Each additional session attended resulted in a 0.57 percentage point increase in the proportion of days attended annually (95% CI: 0.0017–0.0096, p = 0.0051), translating to an estimated 6.8 percentage point increase after 12 sessions (95% CI: 2.04–11.52). Additionally, session completion was associated with a reduction in behavioral incidents, with a decrease of 0.23 incidents per year for each additional session attended (95% CI: −0.49–0.036, p = 0.091), corresponding to an estimated reduction of 2.76 incidents after 12 sessions (95% CI: −5.88–0.43). We performed a sensitivity analysis to assess the impact of controlling for gender on the association between session completion and academic outcomes. In both districts, removing gender as a covariate did not meaningfully affect results (SI Table 2).

### Characteristics associated with session completion

In District A, every one point increase in baseline GPA was associated with attending 1.3 more sessions (95% CI: 0.4–2.19, p = 0.00), highlighting the relationship between program participation and academic engagement. Baseline school attendance showed a positive, though non-significant, association with session completion (coefficient: 5.29, 95% CI: −9.52–20.1). Foster care or homelessness was associated with attending 2.13 fewer sessions on average (coefficient: −2.13, 95% CI: −7.11–2.85). Other factors, including ELL status (coefficient: 0.28, 95% CI: −2.69–3.25) and IEP/504 Plan status (coefficient: −0.29, 95% CI: −2.43–1.85), showed minimal or negligible associations with session completion.

In District B, every 10% increase in baseline school attendance was associated with attending 0.5 more program sessions. Foster care or homelessness was associated with attending 1.08 fewer sessions on average (coefficient: −1.08, 95% CI: −2.84–0.68). Race/ethnicity effects were modest across both districts, with Hispanic students in District B attending 1.01 fewer sessions (coefficient: −1.01, 95% CI: −4.83–2.8) and Black students attending 1.45 additional sessions on average (coefficient: 1.45, 95% CI: −3.26–6.17), compared to White students, though these associations were inconclusive given the wide confidence intervals.

**Table 2:** Baseline demographic and academic characteristics of all students and students referred to the behavioral health intervention in School Districts A and B. The table includes data on academic year, grade level, gender, race, socioeconomic status, IEP/504 enrollment, foster or homeless status, English language learner (ELL) status, attendance proportion, and behavioral incidents. Values for the referred population are reported as counts and percentages, where applicable. Medians and interquartile ranges are provided for attendance, mental health scores, and sessions attended. “NR” indicates data not reported for the overall student population. * Hispanic students are reported as White. ** Socioeconomically disadvantaged refers to students eligible for free or reduced lunch, or whose parents/guardians did not receive a high school diploma.

|  | School District A |  | School District B |  |
| --- | --- | --- | --- | --- |
|  | All students | Referred students | All students | Referred students |
| **Academic year** |  |  |  |  |
| 2021-2022 | 16,000 | 74 (48%) | 21,000 | 105 (44%) |
| 2022-2023 | 16,000 | 79 (52%) | 20,000 | 131 (56%) |
| **Grade** |  |  |  |  |
| 3-5 | NR | 0 (0%) | NR | 93 (39%) |
| 6-8 | NR | 70 (46%) | NR | 120 (51%) |
| 9-12 | NR | 83 (54%) | NR | 23 (9.7%) |
| **Gender** |  |  |  |  |
| Girls | NR | 101 (66%) | NR | 163 (69%) |
| Boys | NR | 52 (34%) | NR | 73 (31%) |
| **Race** |  |  |  |  |
| Asian | 1000 (8%) | 14 (9.2%) | 200 (0.8%) | 6 (2.5%) |
| Black | 200 (1.1%) | 8 (5.2%) | 1000 (5%) | 10 (4.2%) |
| Hispanic | 6000 (40%) | NR* | 17000 (80%) | 203 (86%) |
| White | 7000 (43.6%) | 121 (79%) | 2000 (10%) | 17 (7.2%) |
| Other |  | 10 (6.5%) |  | 0 (0%) |
| **Socioeconomically disadvantaged\*\*** | 5000 (31%) | 70 (46%) | 20,000 (98%) | NR |
| **Has 504 plan or IEP** | NR | 27 (18%) | 2000 (12%) | 8 (3.4%) |
| **Foster care,Homelessness** | 200 (1.4%) | 4 (2.6%) | 800 (4%) | 32 (14%) |
| **English language learner** | 2000 (11%) | 13 (8.5%) | 6000 (27%) | 120 (51%) |
| **Attendance proportion** | NR | 0.95 (0.91, 0.98) | NR | 0.92 (0.84, 0.97) |
| **Behavioral incidents** | NR | NR | NR | 0.00 (0.00, 0.00) |
| **Sessions completed** |  |  |  |  |
| 0 | - | 107 (45%) | - | 61 (40%) |
| 1-3 | - | 34 (14%) | - | 19 (12%) |
| **4-6** | - | 22 (9.3%) | - | 14 (9.2%) |
| **7-11** | - | 45 (19%) | - | 29 (19%) |
| **12** | - | 28 (12%) | - | 30 (20%) |

**Table 4:** This table presents the results of multivariable linear regression analyses examining the relationship between the number of sessions completed and changes in academic (GPA, attendance rates, behavioral incidents) outcome in School Districts A and B. Models were adjusted for key covariates, including English language learner (ELL) status, IEP or 504 Plan status, foster or homeless status, free or reduced price lunch eligibility (District A only), race/ethnicity, and gender. Models were also adjusted for the baseline values of the outcome in each row. Results are presented as coefficients with 95% confidence intervals (CI) and p-values. “-” indicates data not available for a particular outcome or district.

|  | School District A |  |  |  | School District B |  |  |  |
| --- | --- | --- | --- | --- | --- | --- | --- | --- |
| Outcome | N | Change for every additional session attended | Estimated change for attending 12 sessions | P value | N | Change for every additional session attended | Estimated change for attending 12 sessions | P value |
| Yearly GPA | 153 | 0.0024<br>[-0.013-0.018] | 0.0288<br>[-0.156-0.216] | 0.76 | - | - | - | - |
| Behavioral incidents per year | - | - | - | - | 236 | -0.23<br>[-0.49-0.036] | -2.76<br>[-5.88-0.43] | 0.091 |
| Proportion of school days attended | 153 | -0.00047<br>[-0.0021-0.0012] | -0.0056<br>[-0.0252-0.0144] | 0.58 | 236 | 0.0057<br>[0.0017-0.0096] | 0.068<br>[0.0204-0.12] | 0.0051 |

**Table 5:** Multivariable regression results examining the factors associated with session completion in School Districts A and B. Coefficients, 95% confidence intervals (CI), and p-values are reported for variables including English language learner (ELL) status, IEP/504 Plan status, foster care or homeless status, race/ethnicity, gender, attendance rates, GPA, and behavioral incidents. “-” indicates data not available for a particular variable or district. Significant results are highlighted where applicable.

|  | **School District A (N = 153)** |  | **School District B (N = 236)** |  |
| --- | --- | --- | --- | --- |
| **Variable** | **Coefficient** | **P value** | **Coefficient** | **P value** |
| English language learner | 0.28 [-2.69-3.25] | 0.85 | 0.07 [-1.24-1.39] | 0.91 |
| IEP or 504 plan | -0.29 [-2.43-1.85] | 0.79 | 0.66 [-2.63-3.95] | 0.69 |
| Foster care or homeless | -2.13 [-7.11-2.85] | 0.40 | -1.08 [-2.84-0.68] | 0.23 |
| Race: Black | 0.29 [-4.15-4.73] | 0.90 | 1.45 [-3.26-6.17] | 0.54 |
| Race: White | 0.49 [-2.38-3.36] | 0.74 | -0.64 [-4.98-3.69] | 0.77 |
| Race: Hispanic | - | - | -1.01 [-4.83-2.8] | 0.60 |
| Race: Other | -0.32 [-4.53-3.89] | 0.88 | - | - |
| Gender: Male | 0.09 [-1.61-1.79] | 0.91 | 0.59 [-0.72-1.91] | 0.37 |
| Gender: Nonbinary | -5.85 [-15.63-3.93] | 0.24 | - | - |
| Attendance | 5.29 [-9.52-20.1] | 0.48 | 5.1 [0.69-9.5] | 0.02 |
| GPA | 1.3 [0.4-2.19] | 0.00 | - | - |
| Behavioral incidents | - | - | -0.13 [-0.36-0.09] | 0.25 |

## Discussion

We evaluated an individualized 12-week behavioral health intervention tailored for school-age youth, focusing on highly vulnerable populations across two school districts. The intervention, conducted over two academic years, prioritized addressing the mental health needs of students facing significant challenges, including socioeconomic disadvantage, homelessness, and limited English proficiency. In addition, our analysis revealed notable relationships between session completion, sociodemographic characteristics, and academic outcomes across districts. For example, in District B, we found that completing 12 sessions was associated with 12.8 additional school days attended (assuming a 180-day school year) and 2.7 fewer behavioral incidents.

Students in foster care or experiencing homelessness were less likely to attend sessions in both districts, highlighting additional logistical and systemic barriers to participation. These findings underscore the critical importance of tailoring behavioral health interventions to address the unique barriers faced by vulnerable populations, while highlighting the potential of baseline academic and demographic characteristics to inform strategies for enhancing engagement. The unique socioeconomic vulnerabilities of the two districts provide important context for interpreting the study’s findings. In District A, the referred population consisted predominantly of older students (grades 6–12), with a higher representation of socioeconomically disadvantaged students (46%) compared to the overall district population (31%). This group also included a notable proportion of students with Individualized Education Plans (IEPs) or 504 Plans, reflecting the need for targeted mental health support for students with disabilities. These students could have been placed on IEPs for mental health conditions under treatment, although this data was not explicitly collected. Referrals in District A were made directly by school counselors to the behavioral health intervention, suggesting a more straightforward pathway for accessing services,without additional layers of screening or triage, though not all those with a mental health need would have been identified using this method (since no universal screener was applied).

In contrast, District B, with a younger referred population (primarily grades 3–8), demonstrated pronounced socioeconomic and recent immigrant vulnerabilities, with 98% of all students classified as socioeconomically disadvantaged and 51% identified as English language learners. Moreover, 14% of the referred population were either in foster care or experiencing homelessness, underscoring the significant structural barriers to academic success and health faced by many students in this district. Referrals in District B were routed through an intermediary mental health team comprising therapists who provided an additional layer of expertise by screening students for suitability before referral to the intervention. This process may have influenced the composition of the referred population, as more complex cases were often retained for in-house treatment while milder cases were referred out. This more targeted referral base of clients may have been able to uniquely benefit from a telehealth intervention. Additionally, District B implemented a universal mental health assessment conducted by Daybreak Health, a 65-question screener covering eight mental health domains (e.g., anxiety, depression, trauma), which allowed counselors to proactively identify students experiencing mental health symptoms. This systematic screening likely contributed to the district’s ability to more proactively identify and address the needs of many more students with high levels of vulnerability. These contextual differences in referral pathways and population demographics between the two districts highlight the critical role of local systems and resources in shaping access to mental health services. They also underscore the importance of tailoring interventions to meet the unique needs of diverse student populations, particularly those with heightened socioeconomic challenges, from recently immigrated families, in foster care, experiencing homelessness, or other barriers to care (Locke et al., 2017).

These findings align with prior research showing the benefits of school-based mental health interventions on academic outcomes and in reducing disciplinary actions (Grande et al., 2023; Sekhar et al., 2021; Smout et al., 2024; Spiegel et al., 2023; Zhang et al., 2023). However, this study uniquely highlights the heterogeneous outcomes within vulnerable subpopulations. For example, the strong association between baseline GPA and session completion echoes research emphasizing the critical role of academic engagement in the success of behavioral health interventions (Lee et al., 2024). The observation that students experiencing homelessness attended fewer sessions builds on existing literature pointing to logistical and systemic barriers to care faced by transient populations (Winiarski et al., 2020).

This study has several limitations. First, the observational design precludes causal inferences about the impact of teletherapy sessions on academic outcomes. Second, the use of median imputation for missing follow-up GPA and attendance data may have introduced bias, particularly for students with incomplete records. Third, the study’s reliance on GPA and attendance as academic metrics does not capture other potential indicators of academic success, such as standardized test scores or teacher evaluations. Fourth, the generalizability of findings is limited, as the cohorts represent specific geographic and demographic contexts. Finally, heterogeneity in session completion complicates the interpretation of outcomes, particularly for students with fewer sessions completed.

This study found a relationship between completion of teletherapy sessions and improved academic outcomes among students from socioeconomically vulnerable school districts. These findings underscore the importance of understanding the barriers to session completion for subgroups such as students experiencing homelessness, foster care, or those with lower baseline GPAs. Future research should explore tailored strategies to support session completion, including logistical assistance, culturally responsive programming, and enhanced family engagement.

Additionally, longitudinal studies with more robust causal frameworks are needed to better understand the mechanisms linking school-based teletherapy interventions to academic outcomes. By addressing these gaps, we can further refine school-based mental health programs to maximize their impact and equity.

## Data Availability

Aggregate data are available upon reasonable request to the authors.

## Author Contributions

- Conceptualization: AS, GU, CW.
- Data acquisition and processing: CW, AS.
- Data analysis and interpretation: AS, CW, GY.
- Writing and revision: All authors.
- Supervision: AA, DB, GU.

## Supplementary information

**SI Table 1:** Inclusion criteria across Districts A and B.

| Criteria | N students |  |
| --- | --- | --- |
|  | School District A | School District B |
| All referred students | 533 | 385 |
| Has GPA data before, during, and after intervention | 336 | - |
| Has attendance data before, during, and after intervention | 153 | 340 |
| Has behavioral incident data before, during, and after intervention | - | 236 |

**SI Table 2:**
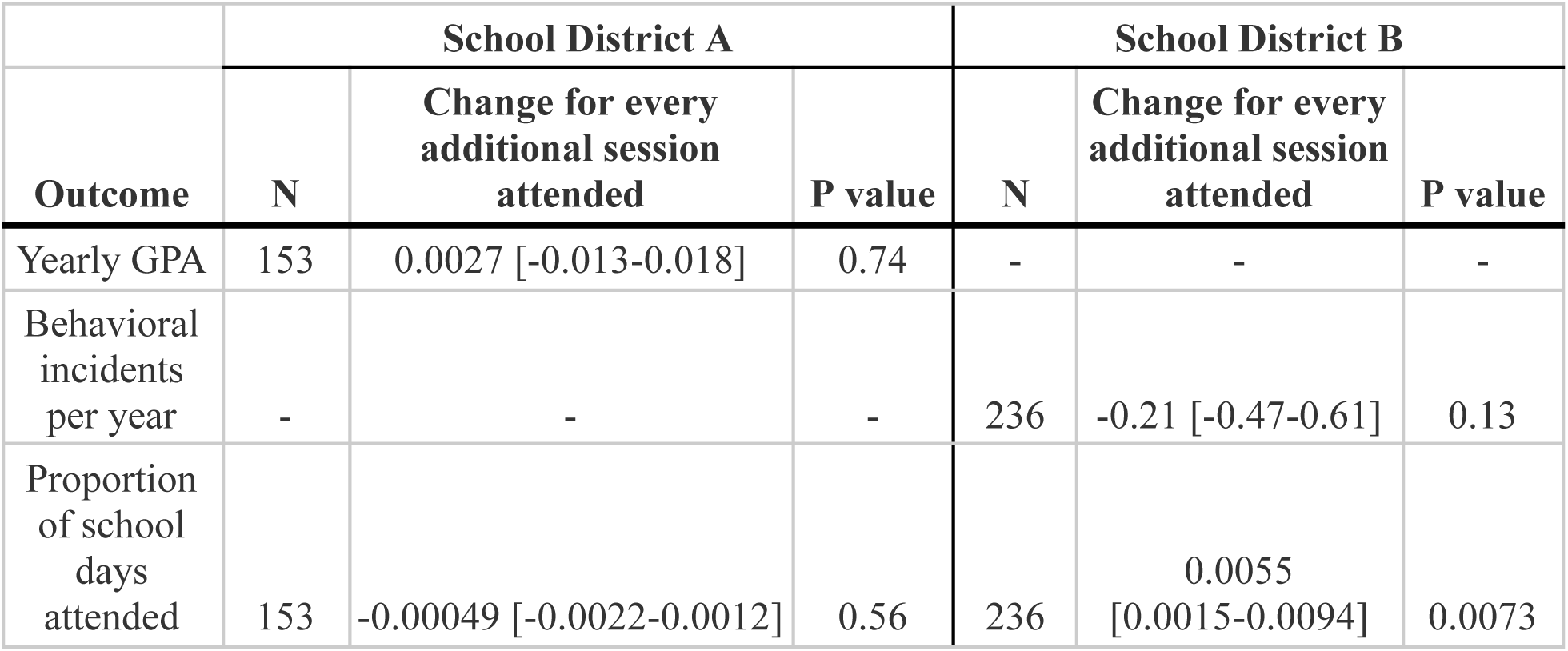
Sensitivity analysis of the relationship between the number of sessions completed and changes in academic outcomes (GPA, attendance rates, behavioral incidents) in School Districts A and B, without controlling for gender. This table presents the results of multivariable linear regression analyses examining the association while controlling for key covariates, including English language learner (ELL) status, IEP or 504 Plan status, foster or homeless status, free or reduced-price lunch eligibility (District A only), race/ethnicity, and baseline values of the outcome in each row. Results are reported as coefficients with 95% confidence intervals (CI) and p-values. “-” indicates data not available for a particular outcome or district.

